# Evaluation of Hemorheological Parameters as Biomarkers of Calcium Metabolism and Insulin Resistance in Postmenopausal Women

**DOI:** 10.1101/2020.11.13.20231050

**Authors:** Paulo L. Farber, Ana Dias, Teresa Freitas, Ana C. Pinho, Diego Viggiano, Carlota Saldanha, Ana S. Silva-Herdade

**Affiliations:** Hospital da Luz, Aveiro, Portugal; Instituto de Medicina Molecular, Faculdade de Medicina, Universidade de Lisboa, Lisboa, Portugal; ESSUA - Escola Superior de Saúde; Universidade de Aveiro, Portugal; Faculdade de Medicina, Universidade de Lisboa, Lisboa, Portugal

**Keywords:** Hemorheology, Calcium metabolism, Osteoporosis, Vitamin D, Insulin resistance, Cardiovascular diseases

## Abstract

**BACKGROUND:** Calcium, vitamin D and insulin resistance are linked to osteoporosis and cardiovascular disease in menopause.

**OBJECTIVE:** Determine if hemorheological parameters related to blood viscosity in microcirculation are linked to calcium metabolism and insulin resistance in menopause.

**METHODS:** 25-Hydroxyvitamin D (25(OH)D)), 1,25-dihydroxyvitamin D3 (1,25(OH)_2_D), Parathyroid Hormone, ionized calcium, Glucose, Insulin and Hemoglobin A1c were measured in blood from 43 volunteers. Red blood cells (RBC) aggregation, RBC deformability and Whole Blood Viscosity were also performed.

**RESULTS:** 25(OH)D showed a positive correlation with RBC deformability 0.60 Pa. Subjects with 25(OH)D ≤ 29.00 ng/mL had lower RBC deformability 0.60 Pa. and higher RBC Aggregation and higher HOMA-IR. Ionized calcium showed a negative correlation with RBC Aggregation. Subjects with ionized calcium ≤1.24 mmol/L showed higher RBC Aggregation. There was a positive correlation between HOMA-IR and RBC Aggregation and HOMA-IR showed a negative correlation with RBC deformability 0.30 Pa. Subjects with HOMA-IR<1.80 showed lower RBC aggregation and higher RBC deformability at 0.30 Pa, 0.60 Pa, 1.20 Pa, 3.0 Pa and 6.0 Pa.

**CONCLUSION:** Low 25(OH)D, low ionized calcium and high HOMA-IR are related to impaired hemorheology in menopause. RBC aggregation and deformability can be used as biomarkers of calcium metabolism and insulin resistance in menopause.

## 1. INTRODUCTION

After menopause, women face an increased risk for several health problems, such as cardiovascular diseases, diabetes and osteoporosis, among others [1]. Cardiovascular diseases are the primary cause of disability and death of these women [2] while osteoporosis fractures are, most likely, the cause for them to have a poor quality of life [3].

Lack of protective hormones such as beta-estradiol, as seen in postmenopausal women, seems to be related with hemorheological (viscoelasticity of blood) and microcirculatory impairment [4–6] and microcirculatory dysfunction are present in both cardiovascular diseases and osteoporosis [7,8].

These facts led us to formulate the hypothesis that hemorheological impairment could be linked for the deleterious effects that happen after the decreasing of hormonal production in women. If confirmed, in addition to contributing to understanding the pathophysiology of menopause, hemorheological parameters could be a target for evaluating treatments.

Hemorheology is the science that studies the deformation and flow of blood [9]. Impaired hemorheology is linked to cerebrovascular diseases [10], cardiovascular diseases [11], coronary heart diseases [12] and osteoporosis [13]. In microcirculation, a decrease of red blood cells (RBC) deformability and an increase of RBC aggregation is known to increase blood viscosity and impair blood flow. Impaired hemorheology interferes in both blood flow and tissue perfusion [14]. Aging related impairment of hemorheological parameters is linked to the aging disease progression [15].

Blood is a non-Newtonian fluid and viscoelasticity of flowing blood is related to hematocrit, plasma viscosity and the behavior of the red blood cells (RBC) suspended in plasma, and dependent of the shear rate. Deformability and aggregation of RBC are characteristics that can interfere with the rheology of blood. The role of RBC in blood flow and viscosity is higher when blood vessels diameter drops than 300 μm [16]. Therefore, RBC aggregation and RBC deformability are critical to blood flow in the microcirculation.

Ovariectomized rats have increased whole blood viscosity (WBV), decreased RBC deformability and increased RBC aggregation [17]. Previous *“ex vivo”* studies showed that β-estradiol decreases RBC aggregation and increases RBC deformability in certain shear stresses [6] and RBC deformability increases with β-estradiol [4]. The results of these studies have shown that WBV, RBC deformability and RBC aggregation may be impaired in postmenopausal women [4,6,17].

In postmenopausal women, and consequently at the stage when cardiovascular risk and osteoporosis rise, two hormones related to calcium homeostasis are often altered, vitamin D and parathyroid hormone (PTH). Serum vitamin D levels decrease with aging and after menopause, and these low levels are related to osteoporosis and an increased incidence of cardiovascular disease [18,19]. PTH levels, on the other hand, can increase in women after menopause, and the elevated values are related to osteoporosis and to cardiovascular diseases, in particular with stroke [20,21].

Vitamin D is fundamental for calcium homeostasis. Vitamin D_3_ is obtained in small amounts from diet and mostly produced by 7-dehydrocholesterol in skin after solar ultraviolet B exposure and hydroxylated into 25-Hydroxyvitamin D (25(OH)D) in liver by 25-hydroxylase. 25(OH)D is stable and is used to measure the vitamin D status. In the kidney, 25(OH)D is converted in the active 1,25-dihydroxyvitamin D3 (1,25(OH)_2_D) by 25(OH)D, 1α hydroxylase (CYP27B1) and 25 hydroxyvitamin D, 24α hydroxylase (CYP24A1). The renal production of 1,25(OH)_2_D is regulated by PTH which are released by parathyroid gland by low levels of ionized calcium. PTH increase bone resorption and calcium absorption in kidney and gastrointestinal calcium absorption [18].

Insulin resistance is also an important risk factor for cardiovascular diseases and can alter bone architecture in postmenopausal women [22,23]. The Homeostasis Model Assessment of Insulin Resistance (HOMA-IR) is one of the best tools for assessment of insulin resistance[24]. Fasting glucose and fasting insulin rise dramatically after menopause and insulin resistance is associated with impaired hemorheology[25,26].

The purpose of this study was to determine if hemorheological parameters are related to calcium, vitamin D, PTH and insulin resistance, and if them can be used as biomarkers in postmenopausal women.

## 2. METHODS

### 2.1. Subjects

Postmenopausal women (n=43) were recruited. The selected subjects had at least one year from menopause, at least 1 ovary and were not on hormone replacement therapy or anticoagulant therapy. In cases of hysterectomized volunteers, subjects with plasma levels lower than 14.2 mIU/mL for luteinizing hormone and lower than19.3 mIU/mL for follicle-stimulating hormone (FSH) [27] were excluded. Blood was collected from antecubital vein after a minimum 8 hours fasting.

### 2.2. Blood tests

Blood samples were collected in three different test tubes from each subject: one with Lithium heparin, one with Ethylenediaminetetraacetic acid (EDTA) and one plain test tube.

Complete blood count, 25-Hydroxyvitamin D (25(OH)D), 1,25-dihydroxyvitamin D3 (1,25(OH)_2_D), PTH, ionized calcium, Fasting Glucose, Fasting Insulin, Hemoglobin A1c (HbA1c), Creatinine, thyroid-stimulating hormone (TSH), LH, FSH, 17-β estradiol and progesterone were measured from blood collection at Synlab lab, *Hospital da Luz do Norte*.

Complete blood count was performed by flow cytometry using Cell-Dyn Sapphire automated hematology analyzer (Abbot, USA). Serum parathyroid hormone, serum insulin, serum 25(OH)D, serum 1,25(OH)_2_D, serum LH, serum FSH, serum 17-β estradiol and serum progesterone were measured by chemiluminescence using Architect I1000sr Analyzer (Abbott, USA). Serum 25(OH)D were measured by chemiluminescence using Architect I2000SR Analyzer (Abbott, USA). Serum 1,25(OH)_2_D were measured by chemiluminescence using Liaison XL Analyzer (DiaSorin, Italy). Serum TSH was measured by chemiluminescence using Architect I2000 Analyzer (Abbott, USA). Fasting glucose and creatinine were measured by colorimetry using Architect c8000 - Chemistry Analyzer (Abbott, USA). Hemoglobin A1c was measured by high performance liquid chromatography using Automated Glycohemoglobin Analyzer HLC-723G7 (Tosoh, Japan). Serum ionized calcium were measured by ion-selective electrodes using ABL800 FLEX blood gas analyzer (Radiometer, Denmark).

The Homeostasis Model Assessment of Insulin Resistance (HOMA-IR) was calculated by the formula Fasting Glucose (mg/dL) x Fasting Insulin (μU/mL)/405 [28].

Relative obesity and Body Mass Index (BMI) was calculated using the formula Weight (Kg)/Height (m) ^2^ [29].

### 2.3. Hemorheological measurements

Hemorheological measurements were performed at Institute of Molecular Medicine, Institute of Biochemistry, Faculty of Medicine, University of Lisbon. Blood samples collected into tubes with lithium heparin (17UI/mL) as an anticoagulant (BD Vacutainer, Portugal) were used for evaluation of hemorheological parameters according with the guidelines for hemorheological laboratory techniques[30].

#### 2.3.1 Erythrocyte aggregation Index (EAI)

RBC aggregation was determined using the MA1 aggregometer (Myrenne GMBH, Germany). The MA1 aggregometer consists of a rotating cone plate chamber which disperses the sample by high shear rate of 600 s^−1^ and a photometer that determines the extent of aggregation. The intensity of light (emitted by a light emitting diode) is measured after transmission through the blood sample. The aggregation will be determined in stasis for 10 and 5 seconds after dispersion of the blood sample.

#### 2.3.2 RBC deformability

RBC deformability was determined by laser diffractometry using a Rheodyn SSD laser diffractometer (Myrenne GMBH, Germany). 20 μL of each blood sample were diluted in 2 mL of dextran solution (Pharmacia, osmolality 0.300, pH 7.4, viscosity 0.24 mPa.s). This suspension was introduced into a measuring chamber formed by two glass disks, one static and other connected through a rotational arm to a synchronized step motor. A 1mV helium-neon laser beam was passed through the blood suspension, and the diffraction pattern analyzed at following shear stress forces values: 0.3, 0.6, 1.2, 3.0, 6.0, 12.0, 30.0, 60.0 Pa. In the static position at rest, the laser diffraction pattern is circular, becoming elliptic as the erythrocyte were deformed by application of an increasing shear stress. Light intensity and the diffraction pattern were measured in two orthogonal axes and the erythrocyte elongation index (EEI) calculated from the length (L) and width (W) by the following formula EEI = 100×(L−W)/(L +W). EEI values are expressed as percentage.

#### 2.3.3. Whole Blood Viscosity (WBV)

WBV was assayed by using Digital Viscometer (Brookfield, EUA), in shear stresses 22,5 s^-1^ and 225 s^-1^, in original hematocrit and in the hematocrit corrected by 45%.

### 2.4. Statistical Analysis

Statistical Analysis was performed with GrafPad Prism 4.0 software. Pearson correlation and linear regression were performed in order to verify the correlation between parameters. According to the results, subjects were split between 2 groups and unpaired, two-tailed t test was performed. Values were considered statistically significant for p < 0.05. Data are presented as Mean ± SD.

### 2.5. Ethical considerations

The study was approved by the local Ethical Committee of *Hospital da Luz de Aveiro* and authorized by Research Committee of *Hospital da Luz*. All volunteers had written explanation about the study and procedures and signed the informed consent. All personal data were protected accordingly with European General Data Protection Regulation.

## 3. RESULTS

43 women were included in this study. 23 subjects had serum intact PTH higher than reference range (10-65 pg/mL)[27]. 20 subjects had prediabetes (HbA1c≥5.7%) and 3 subjects had diabetes (HbA1c≥6.5%) according with American Diabetes Association[31]. 12 subjects had high blood pressure controlled by medicaments. Only 9 subjects did not take any medicines. The BMI ranged from 18.66 to 38.28 kg/m^2^ (26.67±3.975). 15 subjects had normal weight, defined by BMI<25 kg/m^2^, 19 were overweight, defined by BMI between 25 kg/m^2^ and 29.99 kg/m^2^ and 10 were obese, defined by BMI≥30 kg/m^2^ (obesity definition from America College of Cardiology [32]) (**Table 1**). The values obtained for the clinical laboratorial parameters can be observed in **Table 2** and the hemorheological parameters can be observed in **Table 3**.

**Table 1.**
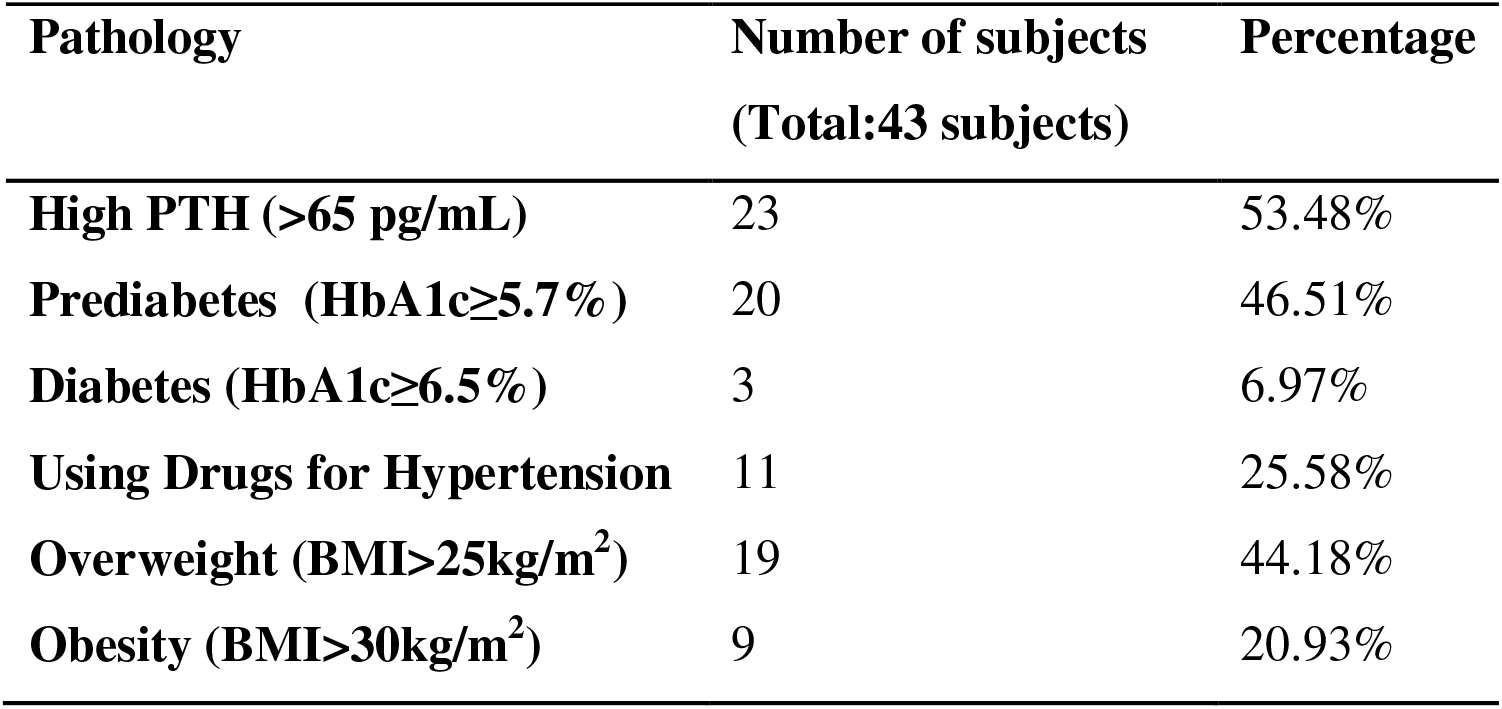
Common pathologies

| **Pathology** | **Number of subjects**<br>**(Total:43 subjects)** | **Percentage** |
| --- | --- | --- |
| **High PTH (>65 pg/mL)** | 23 | 53.48% |
| **Prediabetes (HbA1c≥5.7%)** | 20 | 46.51% |
| **Diabetes (HbA1c≥6.5%)** | 3 | 6.97% |
| **Using Drugs for Hypertension** | 11 | 25.58% |
| **Overweight (BMI>25kg/m<sup>2</sup>)** | 19 | 44.18% |
| **Obesity (BMI>30kg/m<sup>2</sup>)** | 9 | 20.93% |

**Table 2.** Clinical laboratory parameters

| **Parameter** | **Values (mean ± SD)** |
| --- | --- |
| **Glucose (mg/dL)** | 95.63 ± 14.25 |
| **HbA1c (%)** | 5.81± 0.57 |
| **Creatinine (mg/dL)** | 0.75 ± 0.08 |
| **ionized calcium (mmol/L)** | 1.25 ± 0.05 |
| **PTH (pg/mL)** | 73.27 ± 25.72 |
| **Fasting Insulin (μUI/mL)** | 7.32 ± 4.60 |
| **1,25(OH)<sub>2</sub>D (pg/mL)** | 56.69 ± 13.57 |
| **25(OH)D (ng/mL)** | 26.19 ± 11.48 |
| **TSH (mUI/L)** | 1.37 ± 0.67 |
| **LH (mUI/L)** | 27.62 ± 12.43 |
| **FSH (mUI/L)** | 64.01 ± 28.70 |
| **17-β estradiol (pg/mL)** | 6.76 ± 22.50 |
| **Progesterone (ng/mL)** | 0.11 ± 0.07 |

**Table 3.** Values of Hemorheological Parameters namely Hematocrit, RBC Deformability, RBC Aggregation and Whole Blood Viscosity (WBV).

| Parameter | Values (mean ± SD) |
| --- | --- |
| Hematocrit | 41.54 ± 2.88 |
| RBC Aggregation after 5s | 7.95 ± 2.13 |
| RBC Aggregation after 10s | 12.38 ± 2.23 |
| RBC Deformability 0.30 Pa (%) | 3.67 ± 1.45 |
| RBC Deformability 0.60 Pa (%) | 5.94 ± 2.08 |
| RBC Deformability 1.20 Pa (%) | 14.41 ± 2.89 |
| RBC Deformability 3.0 Pa (%) | 27.10 ± 3.41 |
| RBC Deformability 6.0 Pa (%) | 34.29 ± 4.25 |
| RBC Deformability 12.0 Pa (%) | 38.37 ± 5.02 |
| RBC Deformability 30.0 Pa (%) | 40.38 ± 5.55 |
| RBC Deformability 60.0 Pa (%) | 40.01 ± 5.75 |
| WBV 225/s<sup>-1</sup> (mPas) | 4.38 ± 0.23 |
| WBV 22.5/s<sup>-1</sup> (mPas) | 6.80 ± 0.56 |

### 3.1. Age

Age of volunteers ranged from 47 to 70 years. (57.93±5.730). There was no correlation between age and any of the hemorheological parameters. Age had a positive correlation with PTH (Pearson ρ= 0.3337, r^2^= 0.1114, p<0.05) (**Fig. 1A**) and a negative correlation with 1,25(OH)_2_D (Pearson ρ= -0.3487, r^2^= 0.1216, p<0.05) (**Fig. 1B**).

**Figure 1.**
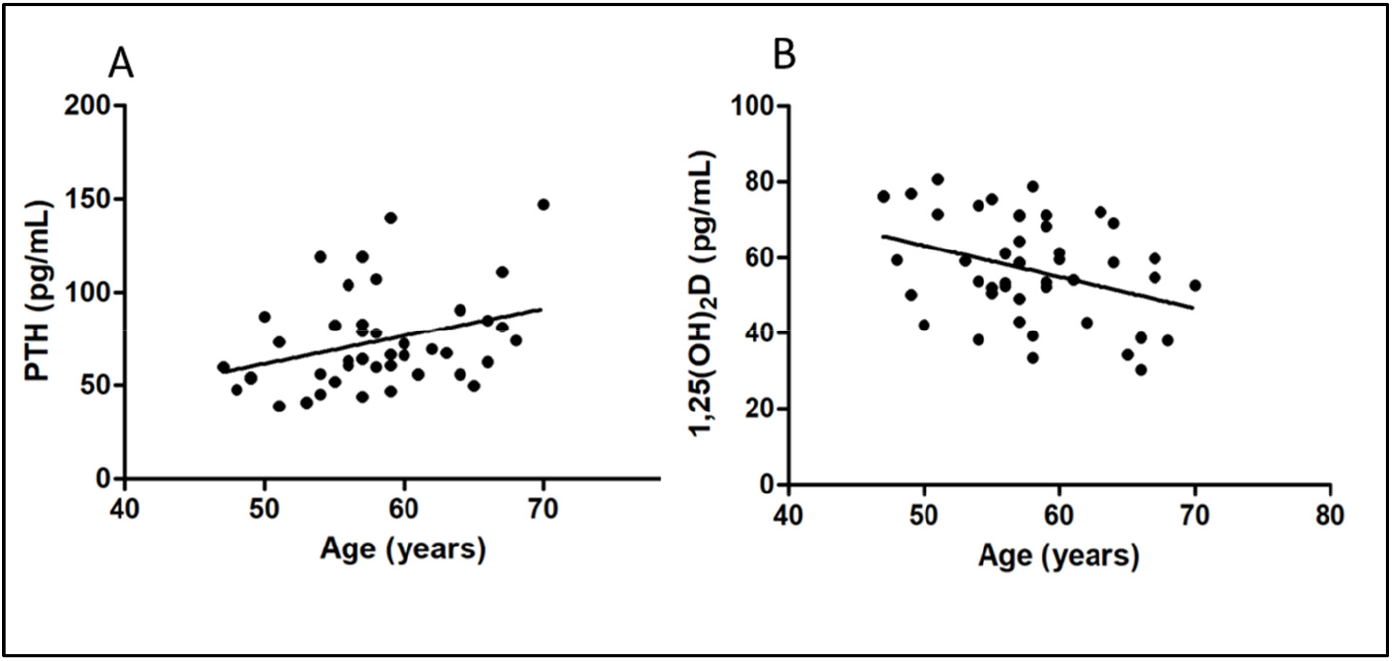
(**A**) PTH (pg/mL) and (**B**) 1,25(OH)_2_D concentrations in function of age (years) in the studied population (N=43). Linear regression showed that PTH levels increases while and 1,25OH_2_D decreases with age progression.

### 3.2. Body Mass Index (BMI)

BMI was positively correlated with PTH (Pearson ρ= 0.3128, r^2^= 0.09787, p<0.05). BMI also was negatively correlated with WBV in both shear stresses 225s^-1^ (Pearson ρ= -0.4377, r^2^= 0.1915, p<0.01) and 22.5s^-1^ (Pearson ρ= -0.3389, r^2^= 0.1149, p<0.05) (**Fig. 2B**) and with Hematocrit (Pearson ρ= -0.3443, r^2^= 0.1186, p<0.05) (**Fig. 2C**).

**Figure 2.**
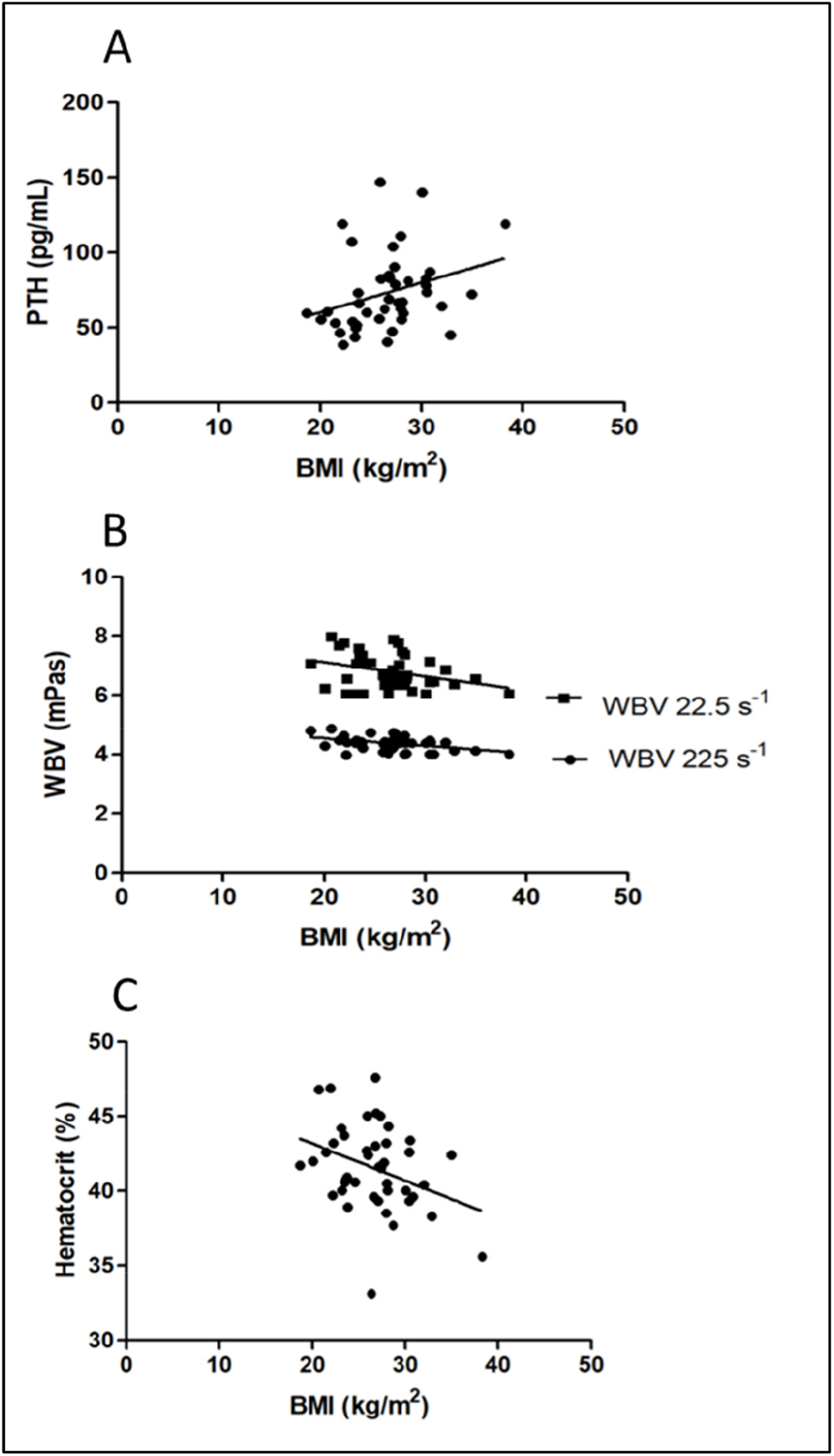
Values of PTH (pg/mL) (**A**), WBV (mPas) (**B)**. and Hematocrit (%) (**C**) in function of BMI (Kg/m^2^). BMI had positive correlation with PTH and negative correlation with 25(OH)D,WBV and Hematocrit.

### 3.3. Hematocrit

Hematocrit was negatively correlated with BMI as showed above (**Fig. 2C**) and had a positively correlated with deformability 0.30 Pa (Pearson ρ= 0.4984, r^2^= 0.2484, p<0.001), 0.60 Pa (Pearson ρ= 0.4574, r^2^= 0.2092, p<0.01), 1.20 Pa (Pearson ρ= 0.6128, r^2^= 0.3755, p< 0.0001), 3.0 Pa (Pearson ρ= 0.4610, r^2^= 0.2125, p<0.01) and 6.0 Pa (Pearson ρ= 0.4113, r^2^= 0.1692, p<0.01). Also there was a positive correlation between hematocrit and WBV, in shear stress 225 s^-1^(Pearson ρ= 0.5160, r^2^= 0.2663, p<0.001) and 22.5 s^-1^(Pearson ρ= 0.5931, r^2^= 0.3518, p<0.001) (**Fig. 3**). There was no correlation between hematocrit and other hemorheological or biochemical parameters.

**Figure 3.**
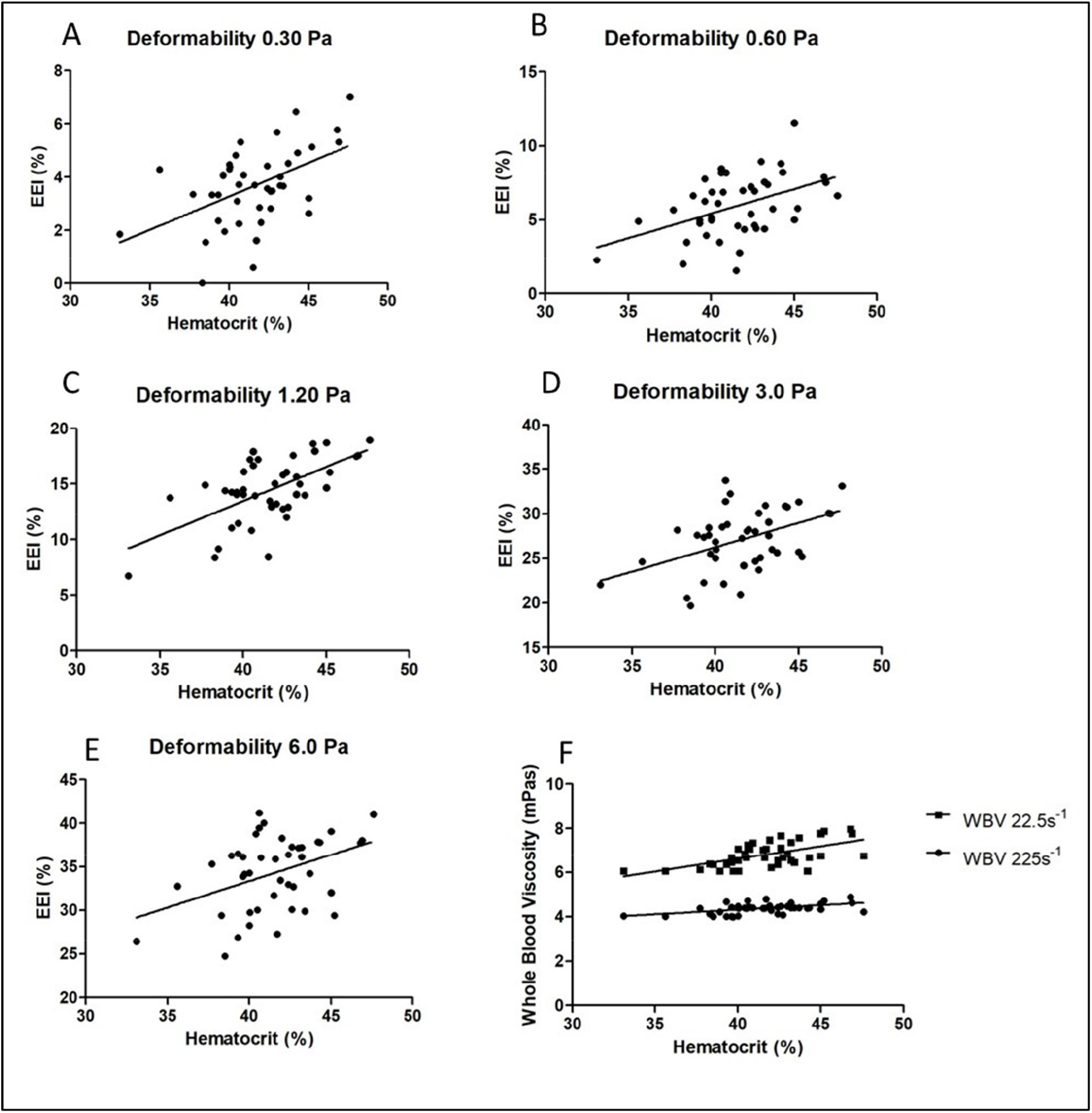
Correlation between hemorheological parameters and hematocrit (%). Hematocrit was positively correlated with RBC deformability in 0.30 Pa (**A**), 0.60 Pa (**B**), 1.20 Pa (**C**), 3.0 Pa (**D**) and 6.0 Pa (**E**). Hematocrit was also positively correlated with WBV in shear stresses 225 s^-1^ and 22.5 s^-1^ (**F**).

### 3.4. PTH, Vitamin D and Calcium

As presented before, PTH showed a positive correlation with age (**Fig. 1A**) and a with BMI (**Fig. 2A**). 25(OH)D showed a negative correlation with Fasting Glucose (Pearson ρ= - 0.3054, r2 0.09329, p<0.05) (**Fig. 4B**). 25(OH)D showed a positive correlation with RBC deformability 0.60 Pa (Pearson ρ= 0.3274, r2 0.1072, p<0.05) (**Fig. 4A**).

**Figure 4.**
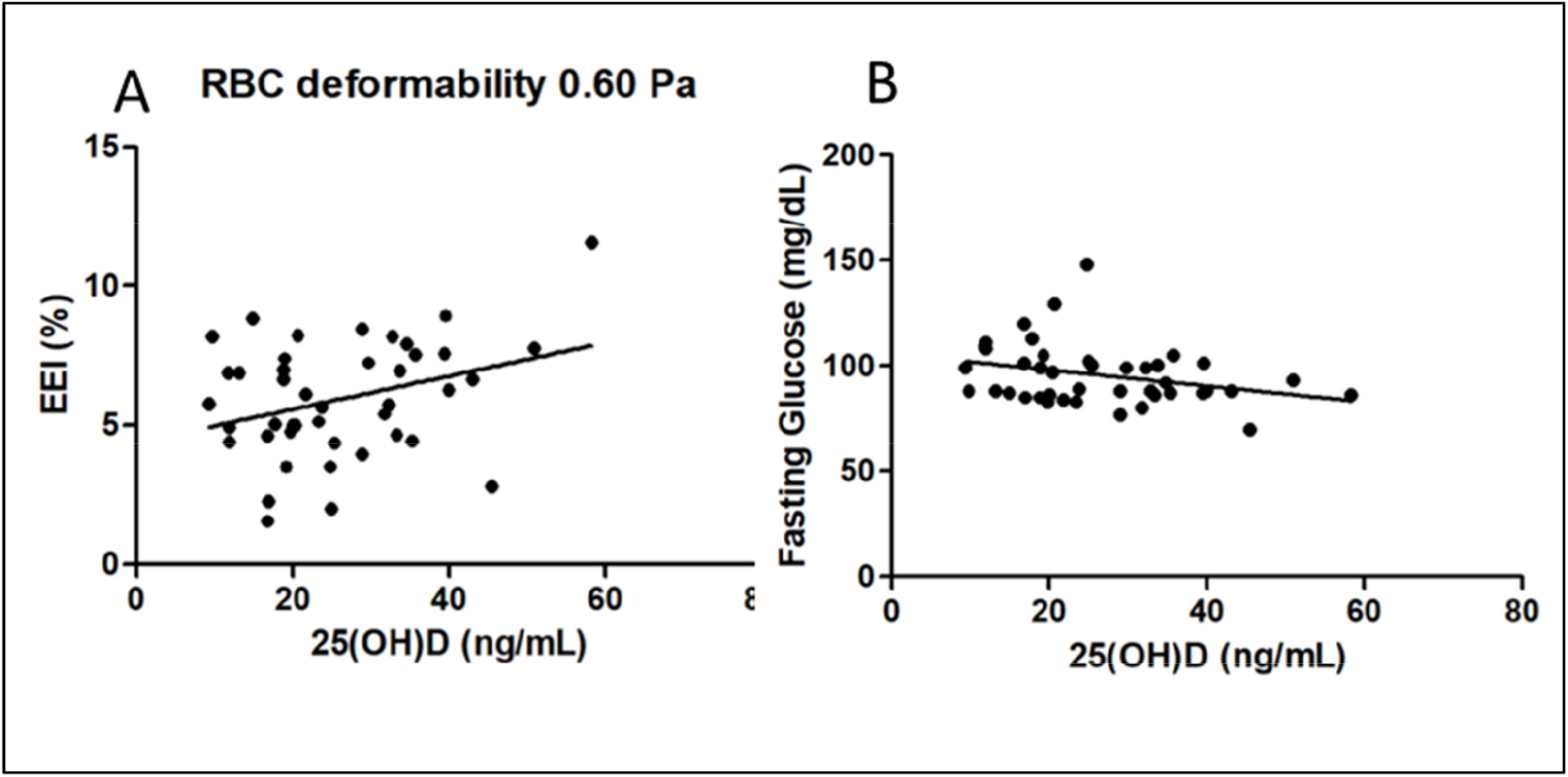
Values of RBC deformability 0.60 Pa (%) (**A**) and Fasting Glucose (mg/dL) (**B**) in function of 25(OH)D concentration. 25(OH)D had a positive correlation with RBC deformability 0.60 Pa and negative correlation with Fasting Glucose.

1,25(OH)_2_D showed a negative correlation with parameters related with insulin resistance: Fasting Glucose (Pearson ρ= -0.3309, r2 0.1095, p<0.05), Fasting Insulin (Pearson ρ= - 0.3313, r2 0.1098 p<0.05), HBA1c (Pearson ρ= -0.3316, r2 0.1100, p<0.05) and HOMA-IR (Pearson ρ= -0.3665, r2 0.1344, p<0.05) (**Fig. 5**).

**Figure 5.**
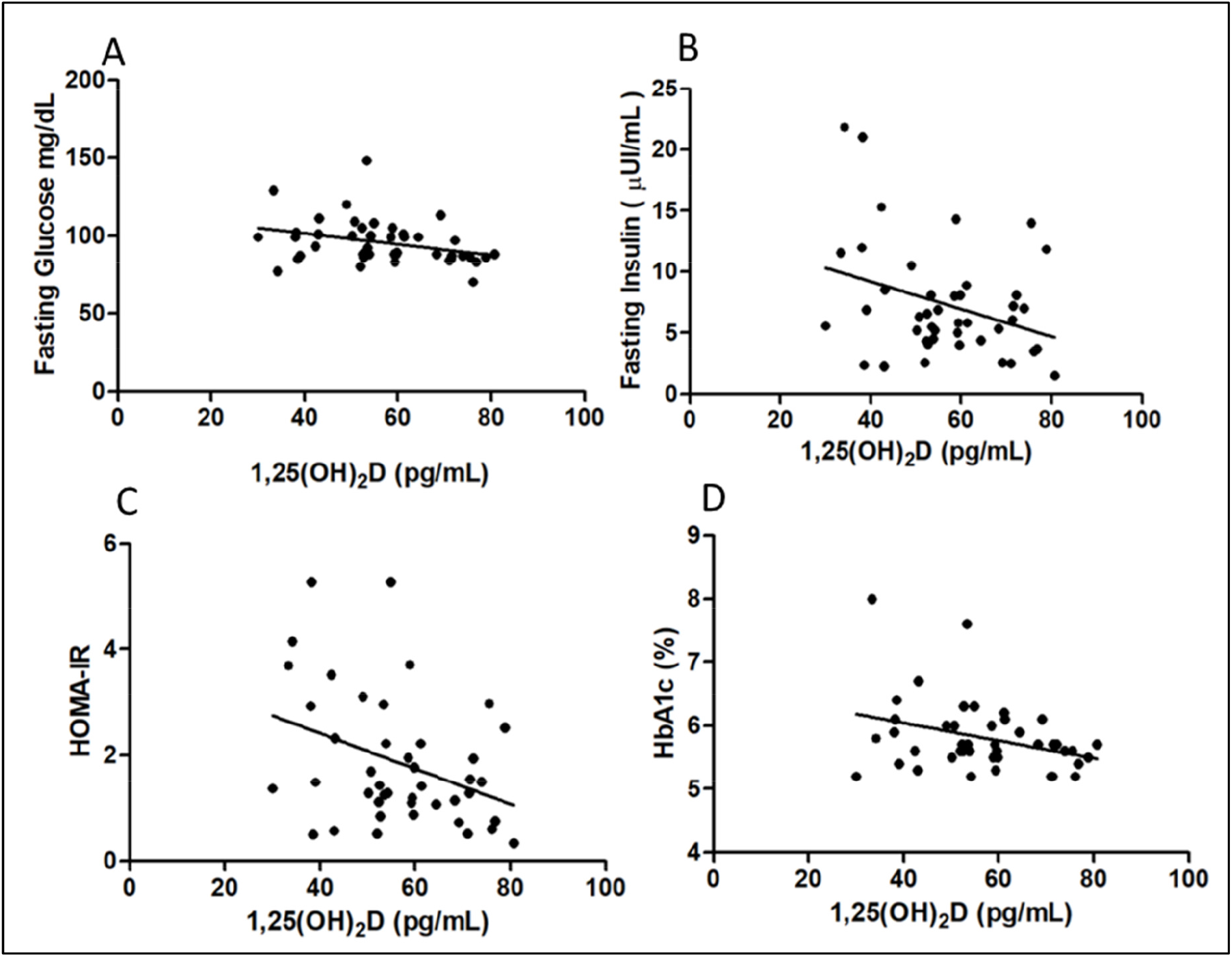
Values of Fasting Glucose (mg/dL) (**A**), Fasting Insulin (µUI/mL) (**B**), HOMA-IR (**C**) and HbA1c (%) (**D**) in function of 1,25(OH)_2_D (pg/mL). 1,25(OH)_2_D was negative correlated with Fasting Glucose, Fasting Insulin, HOMA-IR and HBA1c.

According to the results and for a better analysis of the obtained data, the subjects were divided in two groups, one with 25(OH)D ≤ 29.00 ng/mL (n=27) and other with 25(OH)D > 29.00 ng/mL (n=16). Subjects with lower 25(OH)D had lower RBC deformability 0.60 Pa (5,404 ± 1.954) than the subjects with higher levels of 25(OH)D (6.853 ± 2.037, p<0.05) and higher RBC Aggregation after 5s (8.630 ± 2.054 vs 6,807 ± 1.803, p<0.01). Subjects with 25(OH)D ≤ 29.00 ng/mL also had higher HOMA-IR (2.201±1.322) than those with 25(OH)D > 29.00 ng/mL (1.279±0.8526, p<0.05) (**Fig. 6**).

**Figure 6.**
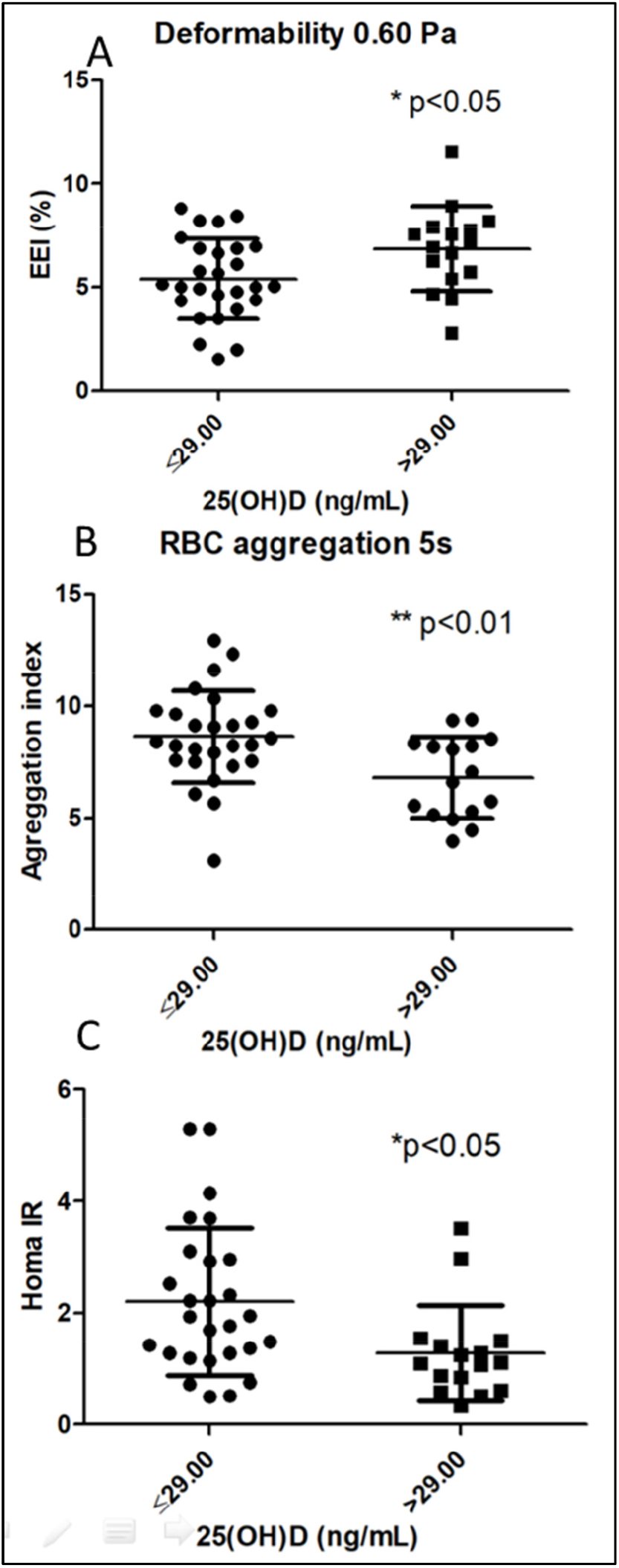
Results of double tailed t test after splitting subjects between 2 groups, according with 25(OH)D levels. Subjects with higher 25(OH)D levels had higher RBC deformability in 0.60 Pa (**A**), lower RBC aggregation after 5 seconds (**B**) and lower HOMA-IR index (**C**).

25(OH)D also showed a positive correlation with ionized calcium. (Pearson ρ= 0.3383, r^2^ = 0.1145, p<0.05). There was no significant correlation between 1,25(OH)_2_D and ionized calcium (data not shown). Subjects with 25(OH)D ≤ 29.00 ng/mL had lower ionized calcium (1.242 mmol/L ± 0.04105) than those with 25(OH)D > 29.00 ng/mL (1. 272 mmol/L ± 0.05369 p<0.05) (**Fig. 7**).

**Figure 7.**
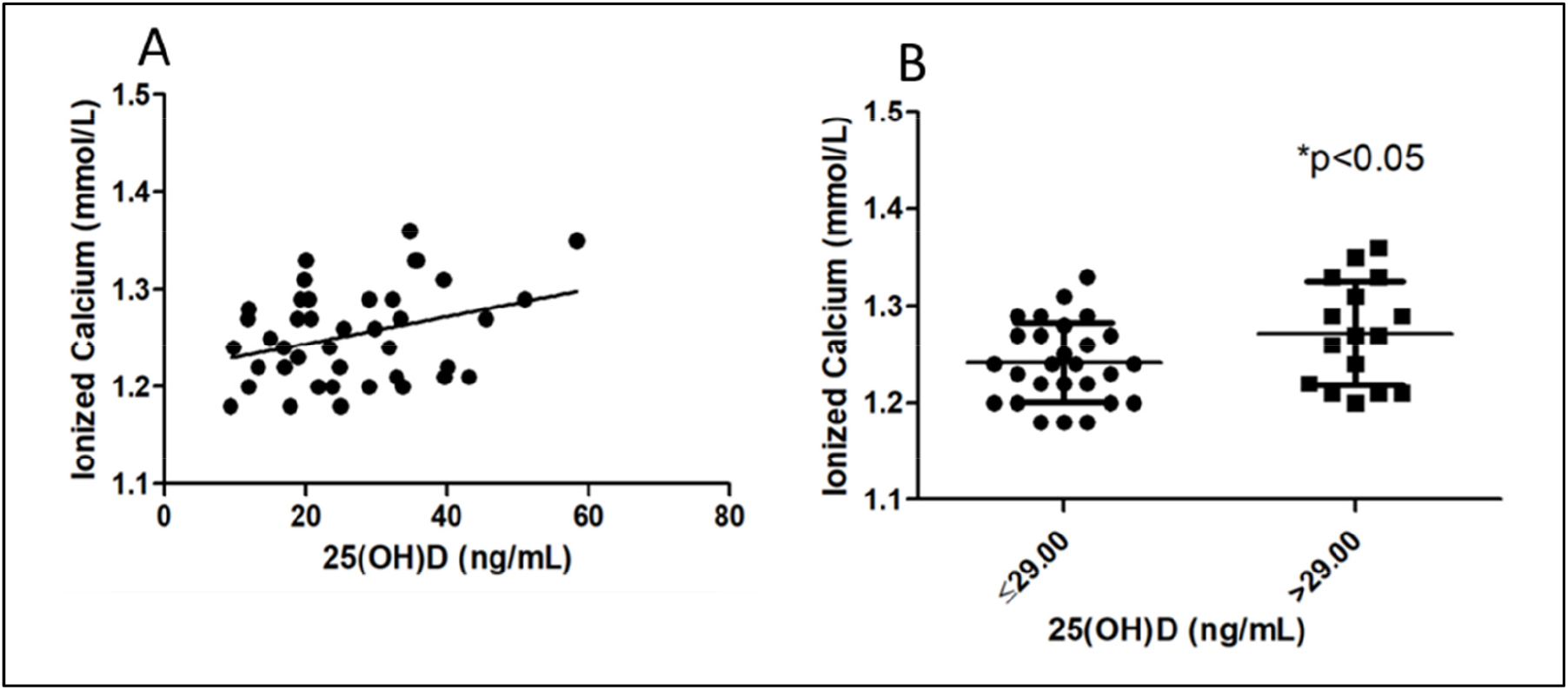
Values of ionized calcium (mmol/L) in function of 25(OH)D (ng/mL) (**A**) and results of double tailed test after splitting 2 groups according with 25(OH)D levels (**B**). Ionized calcium had a positive correlation with 25(OH)D and subjects with lower serum levels of 25(OH)D had lower serum ionized calcium.

Fig. 8A shows that RBC Aggregation after 5s decreases with the increase of ionized calcium (Pearson ρ= -0.4637, r2= 0.2150, p<0.01).

**Figure 8.**
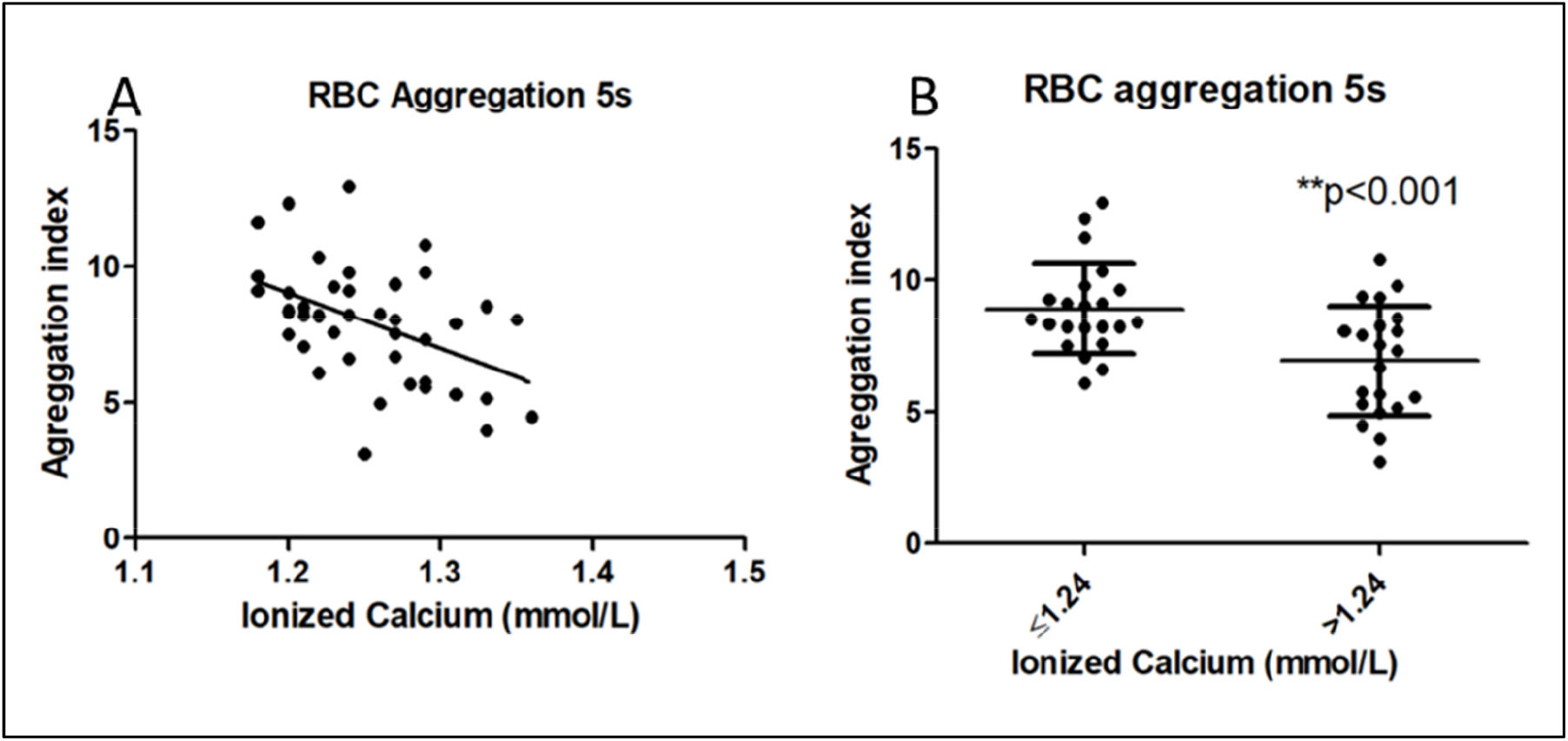
Values of RBC Aggregation after 5s in function of ionized calcium (mmol/L) (**A**) and results of double tailed t test after splitting subjects between 2 groups according to ionized calcium levels. (**B**) Ionized calcium was negatively correlated with RBC aggregation after 5s and subjects with ionized calcium lower than 1.24 mmol/L had higher RBC aggregation after 5s.

According to the results, subjects were split in two groups, one with ionized calcium ≤ 1.24 mmol/L (n=22) and ionized calcium > 1.24 mmol/L (n=21). Subjects with lower ionized calcium levels showed higher RBC Aggregation after 5s (8.921 ± 1.719) than the subjects with higher calcium (6.936 ± 2.091, p<0.01) (**Fig. 8B**).

### 3.5. Insulin Resistance: Fasting glucose, Fasting insulin, HOMA-IR and HbA1c

Concerning the values of HbA1c, no correlations were seen between these values and the hemorheological parameters. Fasting Insulin showed a negative correlation with RBC Deformability 0.30 Pa (Pearson ρ= -0.3346, r^2^= 0.1120, p<0.05) (**Fig. 9**).

**Figure 9.**
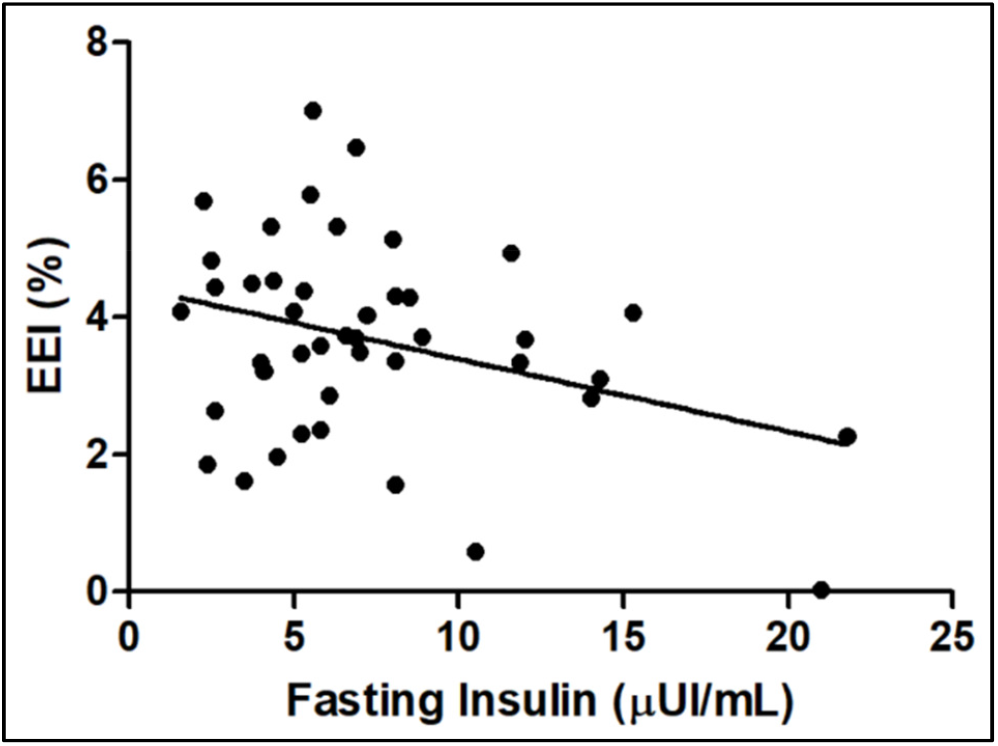
Values of RBC Deformability at 0.30 Pa in function of Fasting Insulin (µ UI/mL). Fasting Insulin was negative correlated with RBC deformability at 0.30 Pa.

There was a positive correlation between HOMA-IR and RBC Aggregation after 5s (Pearson ρ= 0.3263, r2= 0.1065, p<0.05) (**Fig. 10A**) and RBC Aggregation after 10s (Pearson ρ= 0.3039, r2= 0.09, p<0.05) (**Fig. 10B**). There was also a negative correlation between HOMA-IR and RBC Deformability at 0.30 Pa (Pearson ρ= -0,3385, r2= 0,1146, p<0.05) (**Fig. 10C**).

**Figure 10.**
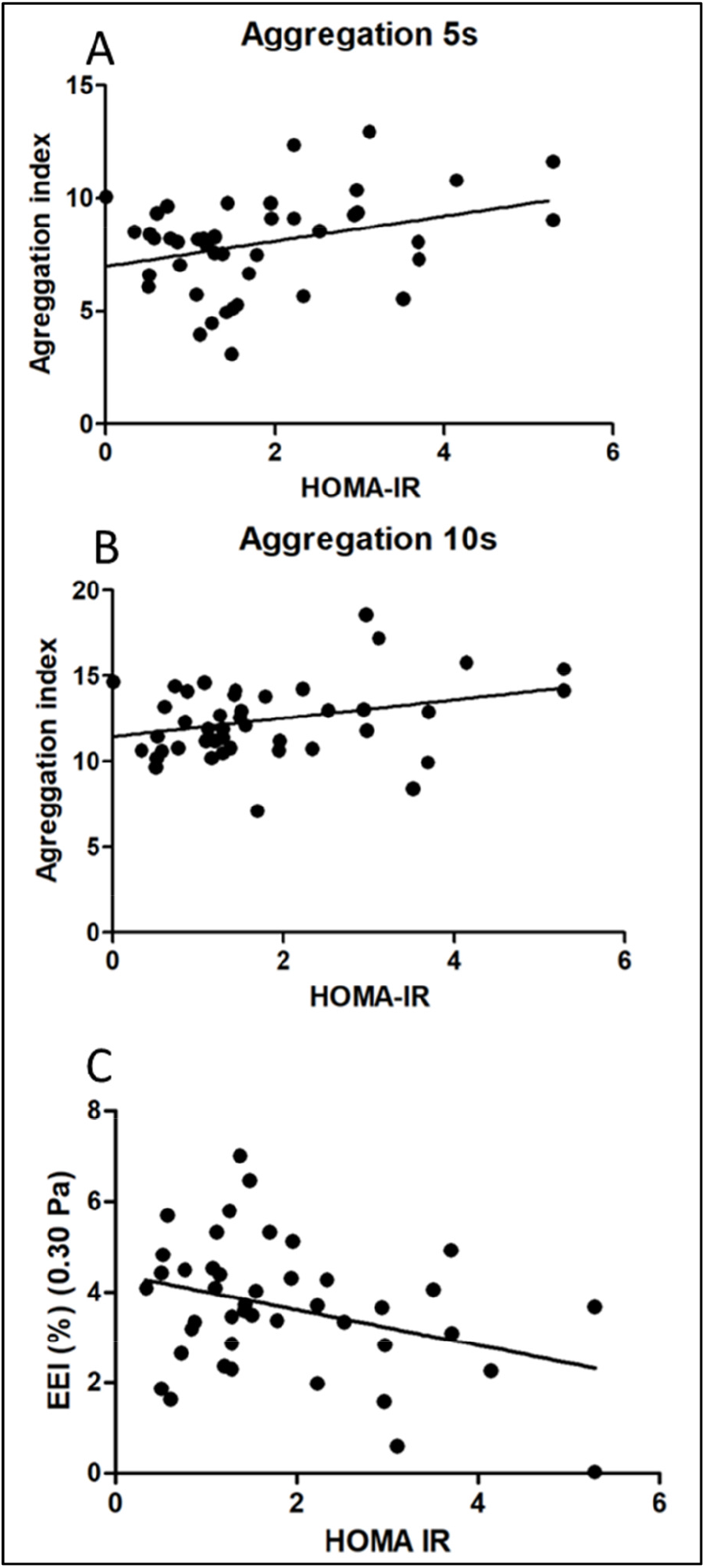
Values of RBC Aggregation and RBC Deformability in funciton of HOMA-IR. There was a positive correlation between HOMA-IR and RBC Aggregation after 5s (**A**) and after 10s (**B**). HOMA-IR also was a negative correlated with RBC Deformability at 0.30 Pa (**C**).

According to the results, subjects were split in two groups, one with HOMA-IR < 1.80 (n=27) and other HOMA-IR > 1.80 (n=16). Higher HOMA-IR had higher RBC aggregation and lower deformability (**Table 4**)

**Table 4.** Results of double tailed t test of hemorheological parameters after splitting subjects between 2 groups according with HOMA-IR. Subjects with HOMA-IR higher than 1.80 (n=16) had higher RBC Aggregation after 5s and lower RBC deformability at 0.30 Pa, 0.60 Pa, 1.20 Pa, 3.0 Pa and 6.0 Pa than subjects with lower HOMA-IR than 1.80 (n=27).

| Hemorheological parameters | HOMA-IR < 1.80 | HOMA-IR > 1.80 | p |
| --- | --- | --- | --- |
| **Aggregation after 5s** | 7,145±1,753 | 9,313±2,078 | <0.01 |
| **Aggregation after 10s** | 11,88±1,734 | 13,22±2,737 | Ns |
| **Deformability 0.30 Pa** | 4,010±1,350 | 3,092±1,472 | <0.05 |
| **Deformability 0.60 Pa** | 6,478±1,944 | 5,039±2,057 | <0.05 |
| **Deformability 1.20 Pa** | 15,18±2,670 | 13,11±2,848 | <0.05 |
| **Deformability 3.0 Pa** | 28,12±3,027 | 25,39±3,428 | <0.01 |
| **Deformability 6.0 Pa** | 35,34±4,095 | 32,51±4,005 | <0.05 |
| **Deformability 12.0 Pa** | 39,43±4,750 | 36,58±5,112 | Ns |
| **Deformability 30.0 Pa** | 41,43±5,256 | 38,61±5,739 | Ns |
| **Deformability 60.0 Pa** | 41,09±5,512 | 38,18±5,856 | Ns |
| **WBV 225/s<sup>-1</sup> Pas** | 4,392±0.2388 | 4,351±0.2223 | Ns |
| **WBV 22.5/s<sup>-1</sup> Pas** | 6,790±0.5260 | 6,765±0.5909 | Ns |

## 4. DISCUSSION

As demonstrated by our results, lower 25(OH)D, lower ionized calcium and higher insulin resistance measured by HOMA-IR were associated with a decrease of RBC deformability and an increase of RBC aggregation. The present study also showed that higher levels of 25(OH)D and 1,25(OH)_2_D are linked to a better glucose metabolism and lower levels correlated with increased insulin resistance.

Previous studies have showed that insulin resistance is directly linked to prediabetes, diabetes and cardiovascular diseases [22]. Serum levels of 25(OH)D are inversely associated with several pathologies, including cardiovascular diseases and disorders of glucose metabolism [33]. Postmenopausal women with type 2 diabetes (T2D) and obesity have lower levels of 25(OH)D[34]. Lower levels of 25(OH)D are also associated with poorer glycemic control in postmenopausal women [35].

This study showed that insulin resistance, the main cause of diabetes [36], measured by HOMA-IR, is associated with RBC aggregation and RBC deformability (**Fig. 10** and **Table 4**). These findings mean that insulin resistance can impair hemorheology and microcirculation. Previous studies have linked diabetes to impaired hemorheology and microcirculation, which may be a cause of cardiovascular problems, nephropathy and retinopathy [11,37–39].

In our study, PTH increases and 1,25(OH)_2_D decreases with aging (**Fig. 1**) and it is known that 1,25(OH)_2_D is protective against age-related osteoporosis [40]. High PTH and low vitamin D levels are associated with osteoporosis in menopause. Patients with high levels or PTH and low levels of 25(OH)D have a higher bone turnover and increased risk of fracture than those with higher levels of 25(OH)D [41].

In our study, higher 25(OH)D was related to lower RBC aggregation and higher RBC deformability (**Fig. 4** and **Fig. 6**). The improvement in hemorheology is linked to the amount of 25(OH)D at the time of collection. It is not possible for us to know whether vitamin D supplementation improve hemorheology, but if it improves, probably the best supplementation protocol should be daily, to ensure a stable vitamin D level. Currently, there are many vitamin D supplementation protocols, and these protocols are usually analyzed together in systematic reviews and meta-analysis [42,43].

The present study suggests that the influence of insulin resistance and vitamin D in muscle and bone in postmenopausal women might be explained by the impairment of microcirculation due to an increase of RBC aggregation and a decrease of RBC deformability. Osteoporosis is more prevalent in T2D and both osteoporosis and T2D are related to an increased cardiovascular risk and a microcirculatory impairment [44–46].

Osteoporotic fracture is an important complication of T2D despite paradoxically higher bone mineral density in these patients [47]. The role of insulin resistance is still controversial and different findings appear in the literature. Some researchers found insulin resistance inversely correlated with bone mineral density while other studies found insulin resistance causing smaller bone size and greater bone mineral density[23,48]. Insulin resistance and lower 25(OH)D levels in postmenopausal women also leads to muscle attenuation and sarcopenia [49,50].

One of the limitations of this study was the fact that 35 out of 43 subjects were taking medicines at the time of the research and the relatively small number of subjects with no medications unable the evaluation of the medicines effect. But besides this, the fact that almost half of the subjects had diabetes or prediabetes and more than half of the subjects had high PTH levels show that the health of postmenopausal women in our community is in a poor condition. Inflammatory parameters such as fibrinogen, C reactive-protein and erythrocyte sedimentation rate were not measured and can interfere with RBC aggregation [51]. Since vitamin D deficiency and insulin resistance are related to inflammatory diseases it is possible that the increase in RBC aggregation can be related to an increase of inflammation [33,52]. Other inflammatory diseases such as rheumatoid arthritis and systemic lupus erythematosus increase RBC aggregation and decrease RBC deformability, and in these patients metabolic syndrome also have higher RBC aggregation and reduced RBC deformability [53], showing that menopause may behaves as an inflammatory disease.

In conclusion, hemorheological parameters such as RBC aggregation and RBC deformability can be considered biomarkers of calcium metabolism and insulin resistance in postmenopausal women. The results also showed a correlation between vitamin D, ionized calcium and insulin resistance. Future studies are needed to clarify the physiopathology of hemorheology in osteoporosis and cardiovascular disease and which intervention can improve hemorheology and microcirculation in postmenopausal women.

## Data Availability

The datasets generated during and/or analysed during the current study are available from the corresponding author on reasonable request.

## ACKNOWLEDGMENTS

We would like acknowledge: Helena Canhão for her help in study design; José Loreto, Ana Abrantes and Luciane Dionisio Manteiga for their help in carrying out the research; the *Hospital da Luz Learning Health* department; the technicians of Synlab lab and Hospital da Luz de Aveiro staff, including nurses, nurses assistants and administrative assistants, without them this research would have been impossible. The main author also would like to acknowledge Frank Hartman whose intellectual contribution to this work was of great significance.

